# BREAst screening Tailored for HEr (BREATHE) - A Study Protocol On Personalised Risk-based Breast Cancer Screening Programme

**DOI:** 10.1101/2021.10.12.21264928

**Authors:** Jenny Liu, Peh Joo Ho, Tricia Hui Ling Tan, Yen Shing Yeoh, Ying Jia Chew, Nur Khaliesah Mohamed Riza, Alexis Jiaying Khng, Su-Ann Goh, Yi Wang, Han Boon Oh, Chi Hui Chin, Sing Cheer Kwek, Zhi Peng Zhang, Desmond Luan Seng Ong, Swee Tian Quek, Chuan Chien Tan, Hwee Lin Wee, Jingmei Li, Philip Tsau Choong Iau, Mikael Hartman

## Abstract

**Background:** Routine mammography screening is currently the standard tool for finding cancers at an early stage, when treatment is most successful. Current breast screening programmes are one-size-fits-all which all women above a certain age threshold are encouraged to participate. However, breast cancer risk varies by individual. The BREAst screening Tailored for HEr (BREATHE) study aims to assess acceptability of a comprehensive risk-based personalised breast screening in Singapore.

**Methods/Design:** Advancing beyond the current age-based screening paradigm, BREATHE integrates both genetic and non-genetic breast cancer risk prediction tools to personalise screening recommendations. BREATHE is a cohort study targeting to recruit ∼3,500 women. The first recruitment visit will include questionnaires and a buccal cheek swab. After receiving a tailored breast cancer risk report, participants will attend an in-person risk review, followed by a final session assessing the acceptability of our risk stratification programme. Risk prediction is based on: a) Gail model (non-genetic), b) mammographic density and recall, c) BOADICEA predictions (breast cancer predisposition genes), and d) breast cancer polygenic risk score.

**Discussion:** For national implementation of personalised risk-based breast screening, exploration of the acceptability within the target populace is critical, in addition to validated predication tools. To our knowledge, this is the first study to implement a comprehensive risk-based mammography screening programme in Asia. The BREATHE study will provide essential data for policy implementation which will transform the health system to deliver a better health and healthcare outcomes.

**Trial registration:** Not applicable.

## Background

Population-based mammography endeavours to reduce mortality via early detection and prompt treatment (1-3). Despite growing evidence of high heterogeneity of breast cancer risk within populations, breast cancer screening programmes commonly recommend starting mammography screening at age 40 or 50 (4). Furthermore, mammographic screening itself has many limitations – over-diagnosis and overtreatment being prime among them (5). While substantially increasing the number of cases of early-stage breast cancer detected, it only marginally reduces the rate at which women present with advanced cancer, as illustrated in the Cochrane reviews (6), Canadian National Breast Screening Study (7) and other studies (8, 9). This has generated international interest in a more risk-stratified approach to the current “one-size-fits all” population screening programmes (10-15).

BreastScreen Singapore (BSS), a nation-wide mammography screening programme in Singapore established in 2002 by the Health Promotion Board, invites women aged 50 to 69 to participate in the early detection of breast cancer. However, only 66% of the target group have reported to ever had a mammogram, and half of them do not adhere to the recommended biennial screening guideline (<30% of the target group were reported to attend mammogram every 2 years) (16). Lukewarm responses to these initiatives have been attributed to a low perception of risk and misperceptions of risk factors and knowledge of breast cancer by women (16-21). A number of studies have since proposed that risk-based screening may improve timeliness of screening. Furthermore, under the current age-based screening paradigm, approximately 30% of diagnosed breast cancer cases in Singapore are women of a younger age than the recommended screening age by the national guidelines (22). The striking difference of ∼10 years in the peak age for breast cancer in between Asian (40 to 50 years) and Western (60 to 70 years) prompts the need to reconsider screening approaches adapted from Western studies in Asia (23). The design and adoption of risk-stratified approach to screening is needful for timely identification and treatment of these high-risk individuals.

Personalised screening enhances an age-based screening paradigm by tailoring screening recommendations to the individual’s risk profile (24). This reduces the rate of false positive results and over-diagnosis in lower risk individuals, thereby providing a more effective method to identify high risk individuals for intervention (25). Currently, to identify high risk individuals, most screening programmes rely primarily on the evaluation of age, family history, clinical and lifestyle factors, and the testing of pathogenic variants in breast cancer predisposition genes (14, 26).

Breast cancer is a multifactorial disease with both genetic and non-genetic risk factors. The Gail model (also known as the Breast Cancer Risk Assessment Tool) was first developed in 1989 for prediction using non-genetic risk factors in Whites, and has since been calibrated and validated for other ethnicities (27). Furthermore, information from the first screen (i.e. mammographic density and false positive status) are indicators of elevated risk (28). The validated Breast and Ovarian Analysis of Disease Incidence and Carrier Estimation Algorithm (BOADICEA) model is able to predict carriership of mutations in known breast cancer genes such as *BRCA1* and *BRCA2* (2, 29-32).

Known pathogenic variants are rare. Due to cost issues, they are usually tested in only high risk individuals (33, 34). Common variants (i.e. single nucleotide polymorphism (SNP)) associated with breast cancer risk have been discovered through genome-wide association studies (35). Individually, these SNPs have minimal effect on risk. However, Mavaddat *et al*. built a polygenic Risk Scores (PRS) – a tally of 313 SNPs – that emerged as a robust means to estimate an individual’s risk of breast cancer (36, 37). The PRS (313 SNPs) is able to reliably predict breast cancer risk, with those in the top centile having a lifetime absolute risk of 32.6% (38). This PRS has been validated in women of Asian descent (38). Despite a growing body of evidence illustrating the utility of PRS in population screening programmes, policy implementation has been low (3). While the Gail model (27), mammographic density (39) and breast cancer predisposition genes (40) have been incorporated into prior risk stratification studies, implementation of PRS is less common (3).

BREATHE is a landmark study aiming to contextualise a personalised, risk-based screening approach to the Asian population (specific aims are listed in **Table 1**). The present study endeavours to explore the acceptability and potential impact on changes in screening behaviour of the BREATHE risk-stratified screening programme as the first step towards policy implementation. With the cost-effectiveness of similar approaches validated (41), it is hoped that BREATHE will greatly enhance resource allocation and patient outcomes in the era of precision medicine.

**Table 1.**
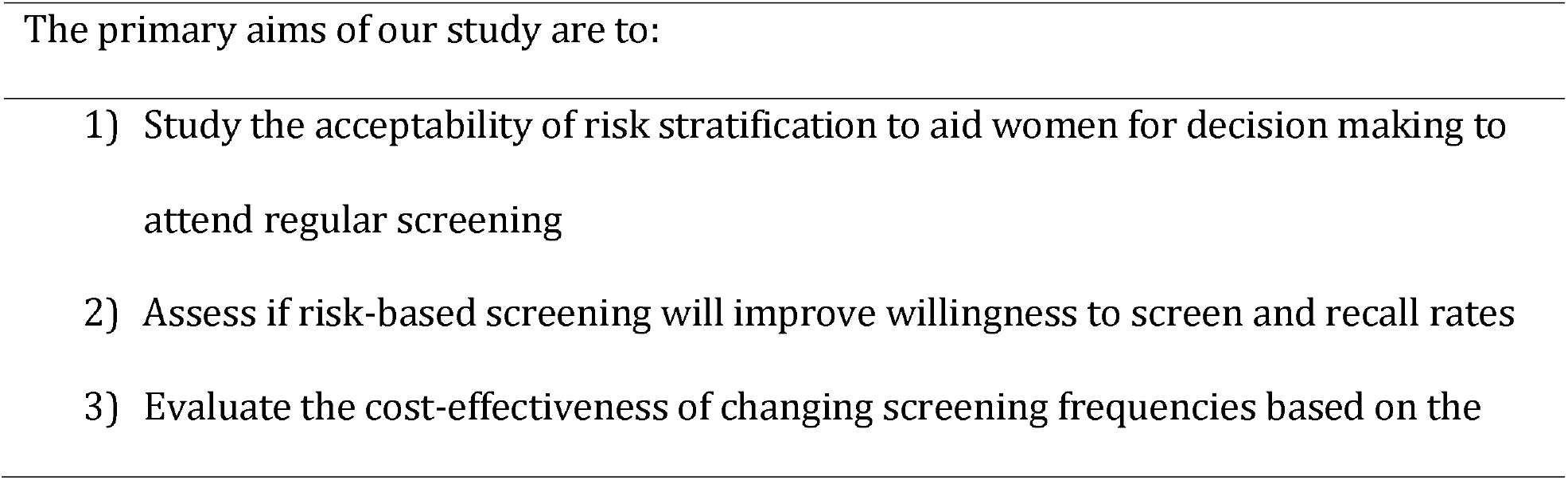
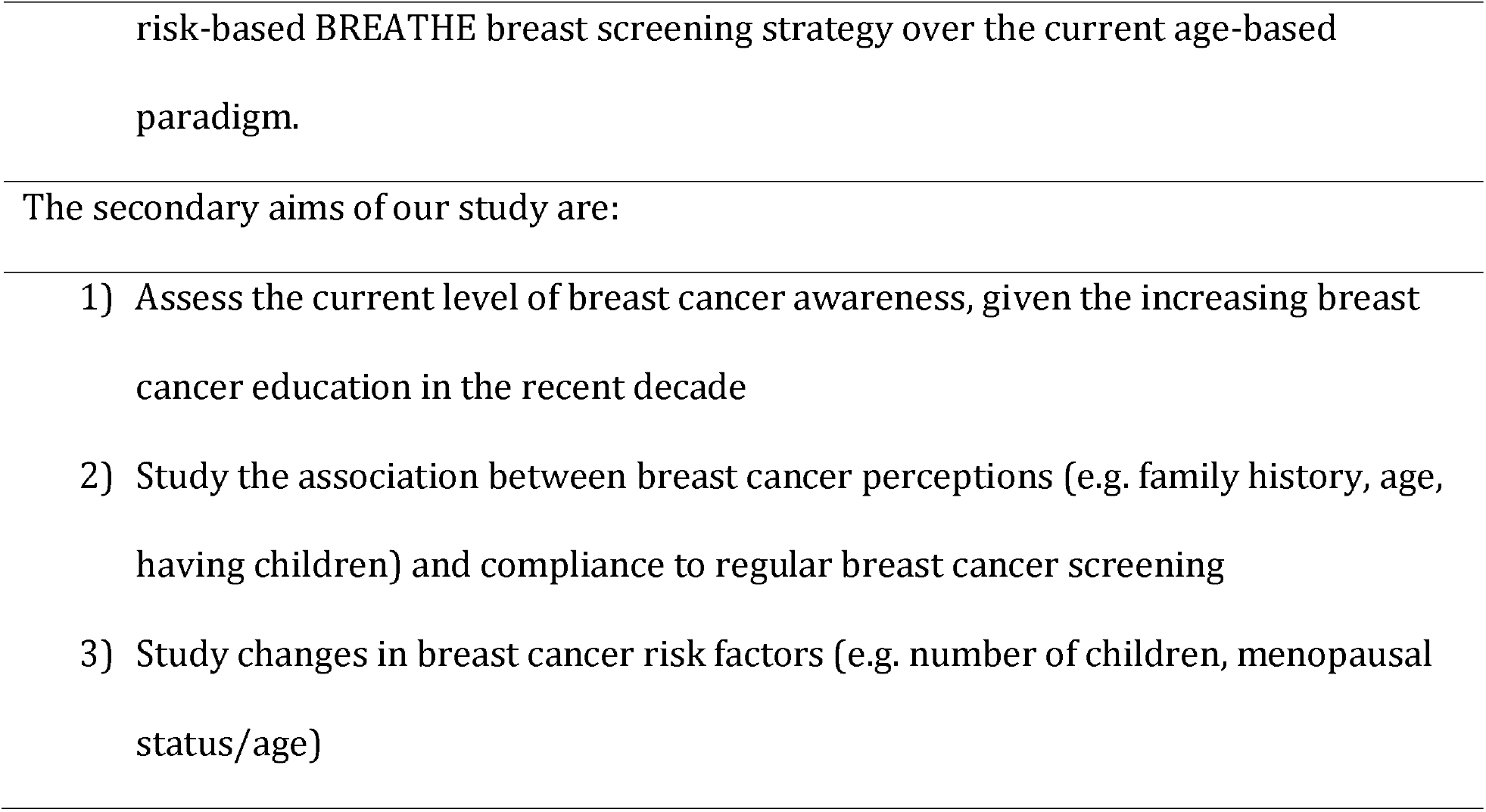
Specific aims of BREAst screening Tailored for HEr (BREATHE).

## Methods/ Design

### Study Design

BREATHE is a prospective multi-centre cohort to study a new modality of breast cancer screening in healthy Singaporean women aged between 35 and 59. We plan to recruit ∼3,500 participants from two hospitals (Ng Teng Fong General Hospital and National University Hospital) and two polyclinics (Bukit Batok Polyclinic and Choa Chu Kang Polyclinic) over a period of two years. To achieve coverage of all age groups of interest, recruitment targets were allocated as such: 20% aged 35 to 39 years; 40% aged 40 to 49 years; and 40% aged 50 to 59 years. The proportion selected was based on the background population in the 2019 population report published on Singapore Department of Statistics. Participants will be on active follow-up for two years. In brief, enrolled participants will be asked (1) to provide a buccal swap for genotyping at study entry and (2) to answer various questionnaires and surveys at study entry and at the two follow-ups (at ∼3 months and ∼2 years after study entry) (**Figure 1**). All surveys are translated to the three major languages used in Singapore: Mandarin, Malay and Tamil.

**Figure 1.**
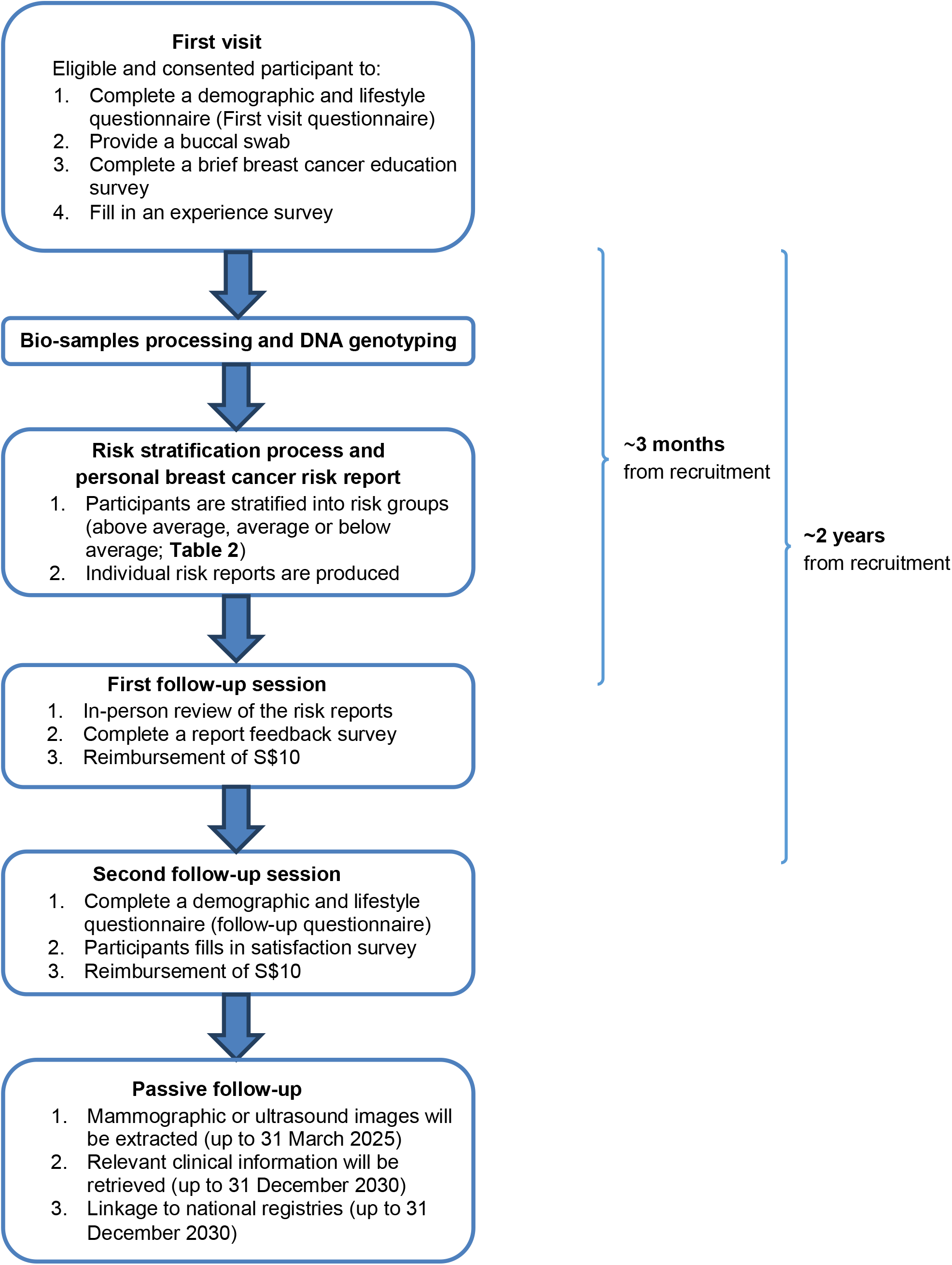
Summary of the recruitment and follow-up process.

### Identification of eligible participants

The study team will identify potential participants (1) through the response to our advertisements on BREATHE (posters, flyers [see Supplementary file 1], and blog.nus.edu.sg/BREATHE) or (2) by approaching them at the participating institutions. The locations include diagnostic departments, women’s clinics and waiting areas of the participating institutions. Responders to our advertisements can either call our hotline, email or fill up an online registration form (see Supplementary file **2**). They will be screened by study team members according to the eligibility criteria. Appointments will be scheduled for eligible participants to visit the participating hospitals or polyclinics for recruitment.

### Eligibility Criteria

Participants must be Singapore Citizens or Permanent Residents, female, aged 35-59 years old. Women who have histologically confirmed diagnosis of any cancer, cognitive impairment which prevent the participant from giving voluntary consent, or are pregnant during recruitment will be excluded.

### First visit

After providing informed consent, participants will be asked to complete a demographic and lifestyle questionnaire and provide a buccal swab sample. A brief education session on breast cancer knowledge and the importance of regular and timely breast self-examination/screening will be self-administered by participants. Depending on their age, the participant will be advised to attend mammography (aged 40 years and above) breast screening. The session will end with a recruitment experience survey.

### First visit questionnaire

Participants will fill in a structured questionnaire detailing various factors associated with the development of breast cancer and related conditions at baseline and over time. These include non-genetic risk factors (demographic, lifestyle, reproductive), past treatments and other environmental factors.

### Buccal swab, DNA extraction and genotyping

Buccal swab (DNA Genotek, ORAcollect-DNA kit) samples will be de-identified and sent in batches (weekly) for deoxyribonucleic acid (DNA) extraction (QIAamp DNA Blood Midi Kit, Part No. 51185). Genotyping will be done (Illumina Global Screening Array [GSA-MD v3.0]) as per manufacturers’ instructions.

DNA extracted from the bio-specimens will be stored in the freezer at -20 degrees Celsius for the duration of our research study. For participants who have agreed to the usage of their bio-specimens for future studies, DNA will be stored after study completion.

### Breast cancer education session

A brief online education session will first assess the screening habits of the participants and their views about breast cancer. Various statements regarding breast cancer will be presented for participants to indicate their agreement. The correct answer and an accompanying explanation are given after every response submitted. The aim is to impart correct information about breast cancer and the importance of regular and timely breast self-examination/screening.

### Mammography screening

Participants may choose to attend screening within the next few months. The study coordinator will assist with setting up appointments for mammography screening with the participating institutions if required. If the participant (aged 40 and above) chooses to attend mammography screen, the study coordinator will seek consent to extract the mammogram image from the service provider (National Healthcare Group Diagnostics (NHGD)). Participants who had a recent mammogram (within one year prior to recruitment) done with NHGD can choose to provide consent for the study to extract the mammogram image.

### Experience Survey

A short survey will be conducted to obtain feedback from the participants on their experience (including any discomfort) with the buccal swab and their initial views about risk-stratified breast cancer screening.

### Risk stratification process and personal breast cancer risk report

Participants will be classified as above average, average or below average risk, based on (1) the Gail model; (2) information from the most recent mammography screening (mammography density and positive recall status); (3) BOADICEA; and (4) the PRS. Participants will first be considered average risk and reclassified as above average or below average based on the criteria in **Table 2**. A risk report will be produced and presented to the participant during the first follow-up session. All participants are recommended to follow current national guidelines (**Table 3**). In the BREATHE programme, women identified to be above average in breast cancer risk are referred to breast specialists at designated study sites, in addition to prevailing guidelines.

**Table 2.**
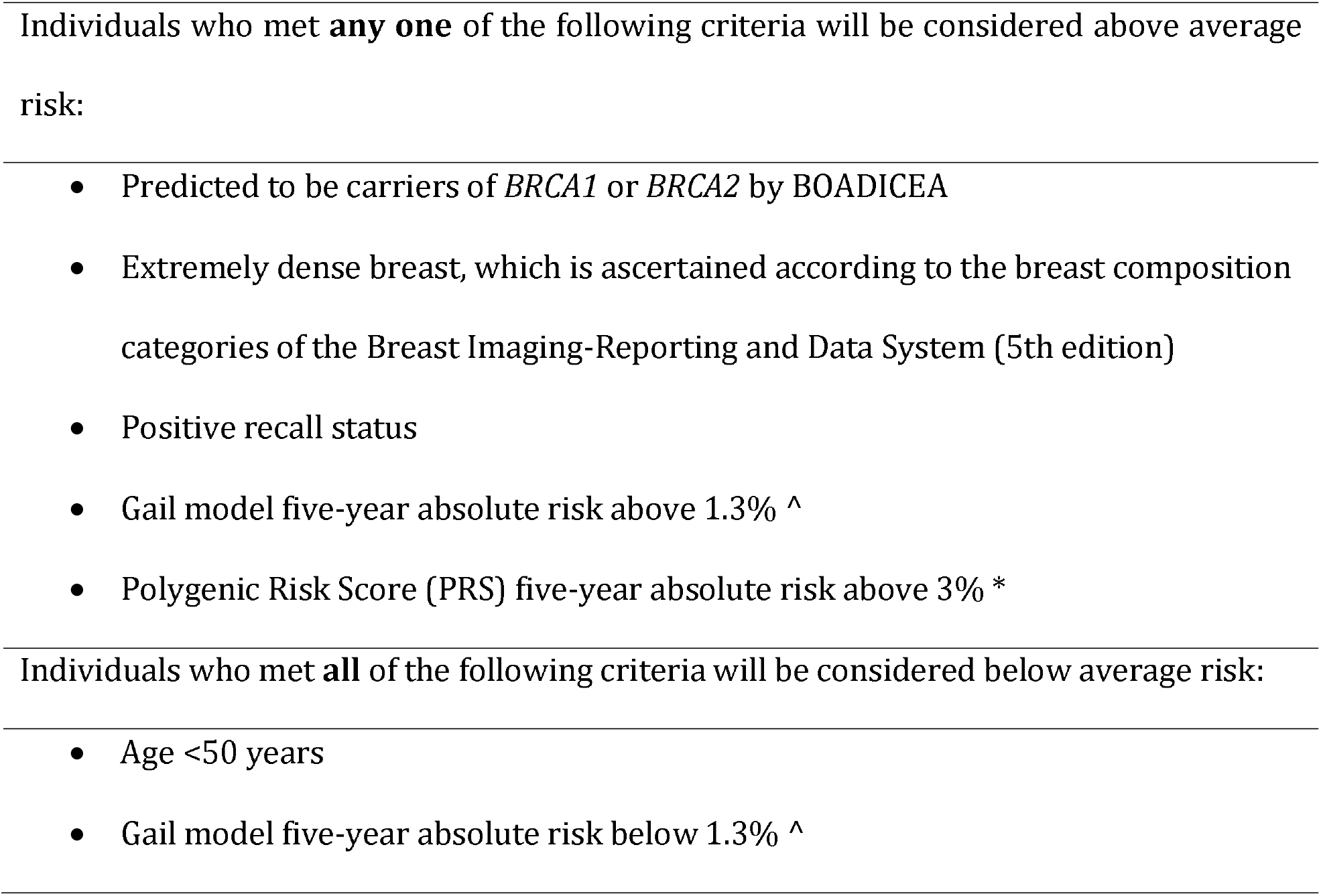
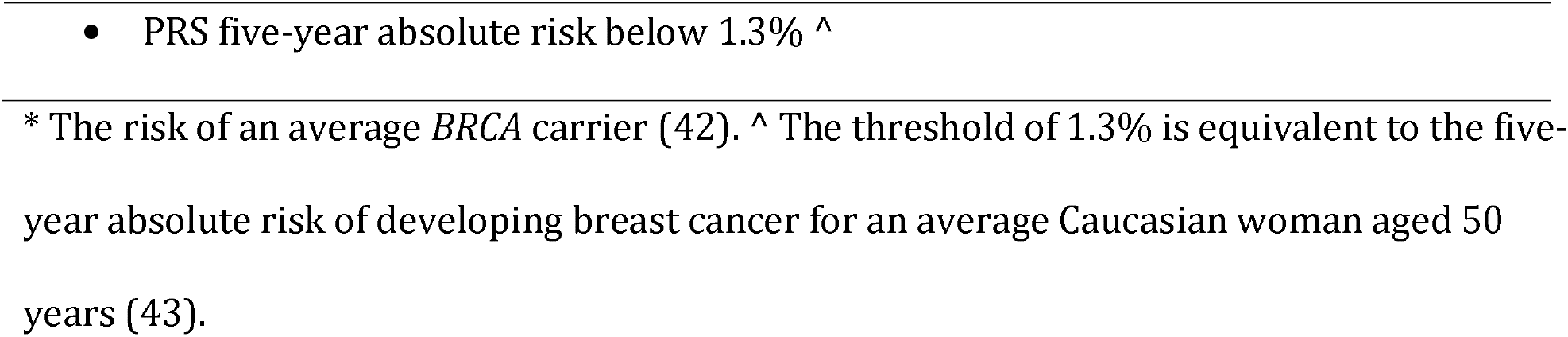
Breast cancer risk reclassification criteria.

**Table 3.** National guidelines for breast cancer screening in Singapore.

| Age groups, years | National guidelines |
| --- | --- |
| 35 to 39 | No recommendation |
| 40 to 49 | Women are to attend yearly mammography screening, if recommended by their doctor. |
| 50 to 59 | Women are to attend mammography screening once every two years. |

### Gail model (non-genetic risk factors)

The Gail Model requires the following breast cancer risk factors from the questionnaire from the first visit: age, age at menarche, age at first live birth, number of previous benign breast biopsies, presence of atypical hyperplasia on biopsy, family history of breast cancer (mother, sisters or daughters), and ethnicity (44, 45). Weights (logistic regression coefficients derived from the Gail model) and attributable risks of Asian-Americans will be used in the calculation of five-year absolute risk based on the Gail model (“Asian.AABCS”, BCRA package in R) (45).

### Information from most recent mammography screening

Mammographic density will be ascertained according to the breast composition categories of the Breast Imaging-Reporting and Data System (5th edition): almost entirely fatty, scattered areas of fibroglandular density, heterogeneously dense or extremely dense. A participant is considered recalled (i.e positive recall status) when she is asked to return for additional confirmatory examination or additional mammography views due to abnormal findings from initial screening.

### Breast and Ovarian Analysis of Disease Incidence and Carrier Estimation Algorithm (BOADICEA) predictions for breast cancer predisposition genes

Carrier probabilities for breast cancer predisposition genes such as *BRCA1* and *BRCA2* will be predicted using the Breast and Ovarian Analysis of Disease Incidence and Carrier Estimation Algorithm (BOADICEA) (40). Briefly, as described by Antoniou *et al* (40), the probability that an individual carries a mutation in *BRCA1*/*BRCA2* or other breast cancer genes based on family history can be computed using Bayes theorem.

### Breast cancer polygenic risk score

PRS is estimated as the weighted sum of effect alleles in 313 SNPs found to be associated with breast cancer; using PLINK (version 3) with the “scoresum” option (46).

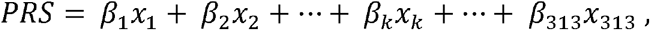

where x_k_ is the dosage of risk allele (0-2) for SNP k, β_k_ is the corresponding weight. The weights of the 313 SNPs for overall breast cancer risk were obtained from are of the overall breast cancer risk published by Mavaddat *et al* (47).

### First follow-up session

The first follow-up session occurs within three months of the recruitment date. This involves an in-person review of the risk reports and ends with a survey on their understanding of the risk report. Participants will be reimbursed S$10 for their time, inconvenience and transportation costs at the end of the first follow-up session.

### Second follow-up session

This is the final in-person follow-up conducted for all participants and occurs approximately two years from date of recruitment. The study coordinator will administer a questionnaire on non-genetic risk factors to capture any changes in participant characteristics since the first visit. The session ends with a satisfaction survey to understand the acceptability of our proposed risk stratification screening programme. Participants will be reimbursed S$10 for their time, inconvenience and transportation costs at the end of the final study visit.

### Passive follow-up

Mammogram images will be extracted if participants have undergone breast screening up to 31 March 2025. In addition, clinical information (e.g. radiology reports, medical conditions, medications and medical reports) related to this study will be retrieved from hospital/polyclinic medical records in accordance to the institutional guidelines, up to 31 December 2030. Clinical information may also be obtained through linkage to nation-wide health-related databases (Singapore Cancer Registry and the Registry of Births and Deaths), and may be done up to 31 December 2030.

### Planned Statistical Analysis

To gauge the acceptability of risk stratification (**Primary Aim 1**) and the current level of breast cancer awareness (**Secondary Aim 1**), descriptive statistics will be performed. Chi-square test for categorical variables and Kruskal-Wallis test for continuous variables will be used for testing differences among risk groups. Post-hoc analysis may be applied for pairwise comparisons.

Logistic regression will be used to study the association between risk perceptions (i.e. risk categories and perceived risk) and follow-up events, which includes actual attendance of breast cancer screening and recall rates (**Primary Aim 2** and **Secondary Aim 2**). Other modelling techniques will be employed dependent on event rates. Adjusted analysis may be done if variability in demographic variables are significant (e.g. conditional logistic models).

To understand potential short-term changes in breast cancer risk factors, paired analysis (e.g. paired-t-test, rank-sum test) between information from the first visit and follow-up will be performed (**Secondary Aim 3**).

Taking the healthcare system perspective, a cost-utility analysis will be conducted to compare BREATHE’s recommendation with the prevailing breast cancer screening guidelines using a Markov model (**Aim 3**). Additional costs associated with breast cancer risk profiling, and changes in healthcare expenditure and health outcomes for different risk groups will be examined. The incremental cost-effectiveness ratio (incremental cost/incremental quality-adjusted life years) will be calculated to understand the cost-effectiveness of BREATH recommendations.

## Discussion

Many previous works have evaluated the validity and discriminatory power of breast cancer risk calculators, alone or in combination (27, 28, 48). In spite of the advances in breast cancer risk prediction, screening recommendations in practice have remained largely unchanged for the past few decades (23). Several large-scale studies conducted in populations of European ancestry, such as KARMA - KARolinska MAmmography Project for Risk Prediction of Breast Cancer (49), PROCAS - Predicting the Risk of Cancer at Screening (50), WISDOM - Women Informed to Screen Depending On Measures of risk (42), are already underway to evaluate the feasibility of implementing risk stratification in breast screening programmes. However, prediction tools should be validated and calibrated to the target population (51). To our knowledge, BREATHE is the first initiative to incorporate risk stratification approaches to enhance the efficacy of existing breast screening protocols in Asia.

Our study leverages on the existing national breast cancer screening programme (BreastScreen Singapore, BSS) (21). Hence mammography service is consistent across all participants. The setup is scalable to include additional hospitals and polyclinics in the future. Singapore is geographically small and convenient for participants to visit breast clinics for recommendations to manage their breast cancer risk. While potential participants can visit multiple hospitals or polyclinics throughout our recruitment period, each individual’s unique National Registration Identity Card number allows us to track them for follow-up. Loss to passive follow-up due to emigration is expected to be minimal for the duration of the study. The BREATHE risk classification is adapted from the established WISDOM Personalized Breast Cancer Screening Trial (42). WISDOM uses a five-year absolute risk threshold of 6% (risk of an average BRCA carrier) for stratification (42). However, it is known that the incidence of breast cancer among Asian women is lower (38, 42). Hence, the BREATHE study uses five-year absolute risk above 3% as a threshold (equivalent to women aged 50 years at the top risk percentile based on PRS in Singapore, data not shown).

BREATHE has some limitations worth noting. Selection bias may arise due to systematic differences between baseline characteristics of responders and non-responders to BREATHE’s advertisements. BREATHE participants may be more health conscious or are already attending breast screening. Such a bias may affect sample representativeness and generalizability of findings. However, the BREATHE study collects information on the study participants (e.g. profession, socio-economic status, highest education attained) and how they found out about the study. This information will allow us to assess the implementation of a risk-based screening approach in this population first, before rolling out the initiative on a larger scale. The BREATHE risk report is based on information available from each participant. For example, if the participant does not participate in or is ineligible for mammography screening, information from first screen will not be in the risk report (participant is assumed to be of average risk). When information is incomplete, breast cancer risk will be underestimated. Barriers to active follow-up two years later are expected. However, the study coordinators will be actively contacting the participants to remind them about the follow-up visit. In addition, the questionnaire is designed such that the participant does not need to be present in-person (conducted electronically or over a phone call).

The aims of BREATHE are aligned with efforts to use personalized health for tailored interventions. For breast cancer screening, multiple studies have supported a risk-stratified approach over the current age-based paradigm due to potentially higher cost-effectiveness and reduced over-diagnosis (13, 24, 52, 53). If BREATHE is successful, women will gain a realistic understanding of their personal risk of breast cancer as well as strategies to reduce their risk, and fewer women will suffer from the anxiety of false positive mammograms and unnecessary biopsies. This work puts Singapore on the world map as a pioneer in integrating state-of-the-art breast cancer risk prediction tools, in particular, breast cancer PRS, in breast cancer screening. This study has real potential to transform breast cancer screening in Singapore.

BREATHE has assembled a multidisciplinary team to build on best practices and emerging data from other risk-based breast cancer screening studies elsewhere. Data-driven and patient-centric value-based care will benefit the healthcare system in many aspects. At the personal level: Women will gain a realistic understanding of their personal breast cancer risk and be empowered to make informed decisions together with their physicians on strategies to manage their risk. At the clinic: The comprehensive risk classification will aid physicians in the conversation on the need for further genetic testing as well as screening and risk reduction strategies. At the population-level: BREATHE generates real-world evidence on how to change the breast cancer screening paradigm to recognize the different needs of individuals. This includes assessment of the organizational readiness, effectiveness, efficiency, resources, costs and cost-effectiveness of implementing a risk-based breast cancer screening approach in Singapore. BREATHE puts Singapore on the world map as one of the pioneers in integrating state-of-the-art risk prediction tools in breast cancer screening, with a real potential to transform the health system to deliver better health and healthcare outcomes.

## Data Availability

Not applicable.

## Supplementary files

File name: Supplementary file 1

File format: PDF

Title of data: Advertisement for BREATHE

Description of data: Advertisements including poster and study brochures for BREATHE.

File name: Supplementary file 2

File format: PDF

Title of data: Online registration form for BREATHE

Description of data: Online registration form for potential participants who are interested in BREATHE study.

## Declarations

### Ethics approval and consent to participate

This study will be conducted in accordance with the ethical principles that have their origin in the Declaration of Helsinki and that are consistent with the Good Clinical Practice and the applicable regulatory requirements, and was approved by the National Healthcare Group Domain Specific Review Board (reference no: 2020/01327). Informed consent will be obtained from the participants.

### Consent for publication

Not applicable.

### Availability of data and materials

Not applicable.

### Authors’ contributions

Conceptualization, H.L.W., J.Li, P.T.C.I. and M.H.; Methodology, J.Liu, P.J.H., S.A.G., Y.W., H.L.W., J.Li, P.T.C.I. and M.H.; Investigation, A.J.K., J.Li, P.T.C.I. and M.H.; Resources, H.B.O., C.H.C., S.C.K., Z.P.Z., D.L.S.O., S.T.Q., C.C.T., J.Li, P.T.C.I. and M.H; Writing—original draft preparation, J.Liu, P.J.H., T.H.L.T. and J.Li; Writing—review and editing, all authors; Supervision, H.B.O., C.H.C., S.C.K., Z.P.Z., D.L.S.O., S.T.Q., C.C.T., J.Li, P.T.C.I. and M.H; Project admin-istration, J.Liu, Y.S.Y., Y.J.C. and N.K.M.R.; Funding acquisition, J.Li and M.H. All authors have read and agreed to the published version of the manuscript.

## Acknowledgements

We want to thank our dedicated research and administrative staff - Ganga Devi D/O Chandrasegran, Hui Min Lau, Pooi Yee Wong, Hui Ling Tan, Kimiie Chia Wei Lin, Nabilah Binte Supiee, Nurfilya Binte Hamdil, Amanda Ong Tse Woon, Jing Jing Hong, Siew Li Tan, Evelyn Low Sok Peng, Marina Mohd, Noor Aisha Binte Mohamed Bahru Ali and Linus Chui for their contributions in the planning and preparation of BREATHE.

