## Supplementary file 1 for "BREAst screening Tailored for HEr (BREATHE) - A Study Protocol On Personalised Risk-based Breast Cancer Screening Programme"

#### I am interested!
#### What should I do now?

**Please register your interest with us either by scanning the QR code**

**or by accessing this link:** [https://tinyurl.com/singaporeBREATHE](https://tinyurl.com/singaporeBREATHE)

#### For more information, please email us or call our study team.



**Research Study**
BREATHE001

### Breast Screening Tailored for Her (BREATHE)

#### Early Detection Saves Lives

**Leading Institution:** Ng Teng Fong General Hospital

**Collaborating Institutions:** National University Hospital; National University Polyclinics

**Supported by:** JurongHealth Fund

*Study Brochure Version 1.6, Dated 16 June 2021*

#### What are the current national guidelines for breast screening?

Based on the national guidelines, women aged 40 to 49 years are recommended to attend mammogram screening yearly, while those aged 50 to 59 years are recommended to attend once every two years. Currently, screening recommendations are age-based.

#### What is the study about?

**BREA**st screening **T**ailored for **HE**r (**BREATHE**) is a research study led by Ng Teng Fong General Hospital (NTFGH) and carried out by National University Health System (NUHS) cluster. **This study aims to explore whether knowing one's risk of having breast cancer will affect their decision to go for regular screening.** The findings from this study may influence a change in current screening programme. Classification of breast cancer risk of an individual is based on genetic risk factors, along with non-genetic factors (i.e. demographics, reproductive and lifestyle risk factors, mammographic density if available). Genetic risk of an individual can be predicted by carrying out a laboratory test performed on DNA, which will be collected via a buccal swab (i.e. cheek swab).

#### Who can join this study?

You will be invited for this research study if you are interested in breast cancer screening and fulfil the following. We are looking for female Singapore Citizens or Permanent Residents aged 35 to 59 years old, with no previous cancer diagnosis and must not be pregnant at the time of recruitment. You must be willing to participate in all research procedures mentioned in the next following sections.

#### Where is the study conducted?

It will be conducted at NTFGH, National University Hospital (NUH), Bukit Batok Polyclinic (BBK) and Choa Chu Kang Polyclinic (CCK).

#### How will I be involved?

**At recruitment**, the research study will be explained to you in detail (approximately 20-30 minutes). Once you have given us your consent, you will be asked to complete a questionnaire (approximately 15 minutes), a buccal swab (approximately 5 minutes), an education session of breast cancer knowledge



(approximately 20-30 minutes), and an experience survey (approximately 5 minutes). These will be performed with a study coordinator.

There will be **2 in-person follow-up sessions** scheduled with you after recruitment. The first follow-up session will be carried out approximately 3 months from your recruitment date, whereby your breast cancer risk report, generated after genetic testing is done on your buccal swab, will be reviewed (approximately 15-20 minutes) and a survey (approximately 10 minutes) will be administered. The final follow-up session will be conducted approximately 2 years from your recruitment date, whereby a questionnaire (approximately 10 minutes) and a survey (approximately 5 minutes) will be administered. You will be reimbursed for your participation upon completion of each follow-up session.

If you have gone and/or will be going for breast screening (a year prior to recruitment and subsequently up to 31 March 2025), we will seek your permission to perform diagnostic images extraction from National Healthcare Group Diagnostics. As part of the study, clinical information from medical records and nationwide health-related databases will also be obtained up to 31 December 2030.

#### Do you accept walk-ins?

No, **we do not accept walk-ins** due to institution's prevailing security and safety measures. Please refer to the last page to register your interest with us.

#### What are the participation benefits?

There is no assurance that you will benefit from participating in this study. Your participation in this study may add to the medical knowledge about breast cancer screening and help to improve the current breast screening method to determine whether knowing one's risk of having breast cancer will affect one's decision to go for regular screening. By participating in this study, you will be receiving your breast cancer risk prediction in the form of a report.

#### Can I withdraw from this study anytime?

Yes, your participation in this research study is voluntary. **You may stop participating in this study at any time**. Withdrawing from this study will not affect your medical care or any benefits which you are entitled to in any of the participating institutions.

### Take time to B.R.E.A.T.H.E

#### BREAst screening Tailored for HEr

#### DID YOU KNOW?

- Breast cancer is **THE MOST COMMON CANCER** among women in Singapore.
- 1 in 13 women is likely to be afflicted by breast cancer[1].
- About 1 in 6 women who suffer from breast cancer are under the age of 45[1].
- The risk of getting breast cancer is different for everyone.

1- Singapore Cancer Registry 50th Anniversary Monograph (1968-2017).

#### EARLY DETECTION SAVES LIVES!

BREATHE is a research study led by Ng Teng Fong General Hospital (NTFGH) and carried out by the National University Health System (NUHS). The participating institutions are NTFGH, National University Hospital (NUH), Bukit Batok Polyclinic (BBK) and Choa Chu Kang Polyclinic (CCK). It aims to explore whether knowing one's risk of having breast cancer will affect their decision to go for regular screening. Research participants will be followed up for approximately 2 years.

**Eligibility criteria:**

- **Singapore Citizen or Permanent Resident**
- **Female, aged 35 - 59 years**
- **No previous cancer diagnosis**
- **Must not be pregnant at the time of recruitment**

**Please register your interest with us by scanning QR code**

or call our study team at 6516 4968. (Monday to Friday, 8.30am to 6pm)

**Leading Institution:** Ng Teng Fong General Hospital

**Collaborating Institutions:** National University Hospital; National University Polyclinics

**Supported by:** JurongHealth Fund

### Take time to B.R.E.A.T.H.E

#### BREAst screening Tailored for HEr

##### DID YOU KNOW?

- Breast cancer is **THE MOST COMMON CANCER** among women in Singapore.
- 1 in 13 women is likely to be afflicted by breast cancer[1].
- About 1 in 6 women who suffer from breast cancer are under the age of 45[1].
- The risk of getting breast cancer is different for everyone.

1- Singapore Cancer Registry 50th Anniversary Monograph (1968-2017).

### EARLY DETECTION SAVES LIVES!

BREATHE is a research study led by Ng Teng Fong General Hospital (NTFGH) and carried out by the National University Health System (NUHS). The participating institutions are NTFGH, National University Hospital (NUH), Bukit Batok Polyclinic (BBK) and Choa Chu Kang Polyclinic (CCK). It aims to explore whether knowing one's risk of having breast cancer will affect their decision to go for regular screening. Research participants will be followed up for approximately 2 years.

**Eligibility criteria:**

- **Singapore Citizen or Permanent Resident**
- **Female, aged 35 - 59 years**
- **No previous cancer diagnosis**
- **Must not be pregnant at the time of recruitment**

**Please register your interest with us by scanning QR code**

**Leading Institution:** Ng Teng Fong General Hospital

**Collaborating Institutions:** National University Hospital; National University Polyclinics

**Supported by:** JurongHealth Fund
