## Supplementary file 2 for "BREAst screening Tailored for HEr (BREATHE) - A Study Protocol On Personalised Risk-based Breast Cancer Screening Programme"

### Take time to B.R.E.A.T.H.E

#### BREAst screening Tailored for HEr

##### DID YOU KNOW?

- Breast cancer is **THE MOST COMMON CANCER** among women in Singapore.
- 1 in 13 women is likely to be afflicted by breast cancer[1].
- About 1 in 6 women who suffer from breast cancer are under the age of 45[1].
- The risk of getting breast cancer is different for everyone.

1- Singapore Cancer Registry 50th Anniversary Monograph (1968-2017).

### EARLY DETECTION SAVES LIVES!

BREATHE is a research study led by Ng Teng Fong General Hospital (NTFGH) and carried out by the National University Health System (NUHS). The participating institutions are NTFGH, National University Hospital (NUH), Bukit Batok Polyclinic (BBK) and Choa Chu Kang Polyclinic (CCK). It aims to explore whether knowing one's risk of having breast cancer will affect their decision to go for regular screening. Research participants will be followed up for approximately 2 years.

**Eligibility criteria:**

- **Singapore Citizen or Permanent Resident**
- **Female, aged 35 - 59 years**
- **No previous cancer diagnosis**
- **Must not be pregnant at the time of recruitment**

**Leading Institution:** Ng Teng Fong General Hospital

**Collaborating Institutions:** National University Hospital; National University Polyclinics

**Supported by:** JurongHealth Fund

BREATHE Poster_ Portrait Version 1.4, Dated 16 June 2021

#### BREATHE Study Information

BREAst screening Tailored for HEr (BREATHE) is a research study led by Ng Teng Fong General Hospital (NTFGH) and carried out by National University Health System (NUHS) cluster. This study aims to explore whether knowing one's risk of having breast cancer will affect their decision to go for regular screening. The findings from this study may influence a change in current screening programme. Classification of breast cancer risk of an individual is based on genetic risk factors, along with non-genetic factors (i.e. demographics, reproductive and lifestyle risk factors, mammographic density if available). Genetic risk of an individual can be predicted by carrying out a laboratory test performed on DNA, which will be collected via a buccal swab.

You will be invited for this research study if you are interested in breast cancer screening and fulfil the following. We are looking for female Singapore Citizens or Permanent Residents aged 35 to 59 years old, with no previous cancer diagnosis and must not be pregnant at the time of recruitment.

Please register your interest by completing the form below. If you have any further queries, please contact our study team for more information at 6516 4968 (Monday-Friday, 8:30 am to 6 pm) or

Collection, use and disclosure of your personal data shall be in accordance with our privacy policy which is available at https://www.nuhs.edu.sg/Pages/Personal-Data-Protection-Act.aspx.

#### To register

Complete the following form. Our study team will be in touch with you in 2-3 working days. Please fill in the required fields*

**1. By submitting this form, I hereby authorise, agree and consent to allow National University Hospital (S) Pte Ltd to collect, use, disclose and/or process my personal data for the purpose of processing, handling and managing my participation in the study stated herein.***

☐ Agree

**2. Name***

**3. Mobile Number***

**4. Email** (optional)

**5. Are you a Singaporean Citizen or Permanent Resident?***

☐ NO ☐ YES

**6. Are you a female aged between 35 - 59 years old?***

☐ NO ☐ YES

**7. Are you currently pregnant?***

☐ NO ☐ YES

**8. Have you had any history of cancer?***

☐NO ☐YES

**9. Please select your preferred participating institution:***

☐Ng Teng Fong General Hospital (NTFGH)
☐National University Hospital (NUH)
☐National University Polyclinic - Bukit Batok
☐National University Polyclinic - Choa Chu Kang

**11. Remarks** (optional)
